# High prevalence of putative invasive pulmonary aspergillosis in critically ill COVID-19 patients

**DOI:** 10.1101/2020.04.21.20064915

**Authors:** Alexandre Alanio, Sarah Dellière, Sofiane Fodil, Stéphane Bretagne, Bruno Mégarbane

**Author notes:** Corresponding author: Dr Alexandre Alanio. Saint Louis Hospital, 1 avenue Claude Vellefaux, 75475 Paris CEDEX 10,.

## Abstract

We are currently facing a frightening increase in COVID-19 patients admitted to the ICU. Aiming at screening for fungal secondary pneumonia, we collected the data of our first 27 ICU patients, who underwent bronchoalveolar lavage or bronchial aspirates. We classified the patients based on the recently published study on invasive aspergillosis in influenza patients (Schauwvlinghe et al., 2018.) and found 33% of our COVID-19 patients with putative invasive pulmonary aspergillosis. Observing such a high prevalence in COVID-infected patients was somehow unexpected since the 30% prevalence of invasive aspergillosis in influenza patients has been attributed to the action of oseltamivir on anti-Aspergillus immunity. Almost all critically ill COVID-19 patients develop ARDS and are likely to receive high-dose steroids or immunomodulatory therapies to prevent worsening as suggested by reports from China. In the COVID-19 patients with putative invasive aspergillosis, antifungal prophylactic therapy may be questioned to avoid increased lung inflammation that may compromise the outcome. This issue remains to be addressed in future clinical trials. We are strongly convinced that testing deep lung specimens for Aspergillus in severe COVID-19 patients should be recommended.

---

About 5% of coronavirus disease 2019 (COVID-19) patients require intensive care unit (ICU) management.^1^ In the ICU, these patients are at high risk of developing secondary infections including invasive pulmonary aspergillosis (IPA).^2^ First reported with H1N1 influenza, IPA represents a frequent (20-30%) and early-onset complication (median, 3 days post-ICU admission) in critically ill influenza patients leading to enhanced illness severity and mortality rate (40-60%).^3,4^ Interestingly, most influenza-associated IPA cases were observed in non-immunocompromised patients, thus questioning the applicability of the EORTC-MSG consensus criteria used largely to define aspergillosis in immunocompromised patients.^5^ Therefore, in the ICU patients without the usual risk factors, a clinical algorithm to discriminate *Aspergillus* colonization from putative IPA was recently developed based on mycological criteria combining visualization and culture of *Aspergillus* from respiratory specimens and galactomannan detection in the bronchoalveolar lavage (BAL) and serum.^4,6^ Paralleling what has been reported in influenza patients, we designed this prospective observational bi-center study to investigate the risk of IPA in critically ill COVID-19 patients, especially since these patients were likely to receive immunomodulatory therapies. The patients were classified using the influenza-associated IPA criteria in the ICU4 completed with beta-D-glucan measurement and quantitative real-time PCR (qPCR) in BAL. Putative IPA was considered if one of the following conditions was met, i.e. 1-presence of *Aspergillus fumigatus* in culture; 2-BAL galactomannan index >0.8 AND beta-D-glucan >80 pg/mL; 3-*Aspergillus fumigatus* qPCR with quantification cycle <35 in pulmonary specimens;^7^ and/or 4-serum beta-D-glucan >80pg/mL AND serum galactomannan index >0.5. Of note, direct examination of respiratory specimens was not performed to avoid operator contamination. Twenty-seven successive mechanically ventilated COVID-19 patients with pneumonia admitted between March 16^th^ and 28^th^ to our two ICUs were included (Table 1). Respiratory specimens (20 BALs and 7 bronchial aspirations) were obtained on day 3 [1-6] post-intubation. Putative IPA was diagnosed in 9/27 patients (33%) including six patients validating ≥2 mycological criteria and three patients with only *Aspergillus fumigatus* identification in the respiratory specimen culture. History of hypertension was significantly more frequently reported in the patients with putative IPA than patients without aspergillosis (*p*=0.04). No other significant differences were observed in terms of age, invasive aspergillosis EORTC-MSG risk factors, time between onset of symptoms and intubation and times between onset of symptoms or intubation and *Aspergillus* respiratory specimen collection, ARDS severity, clinical and laboratory data, non-COVID CT-scan images and steroid administration. No other immunosuppressant drug was administered in the ICU. Specific anti-*Aspergillus* therapy was initiated in only one of the nine patients with putative IPA. In this patient initially receiving caspofungin to treat concomitant invasive blood *Candida glabrata* infection, antifungal treatment was switched to voriconazole. No significant increase in fatality rate was observed in the patients with putative IPA (3/9 *versus* 3/18, *p*=0.4), although so far, 2/9 and 8/18 patients are still intubated in each group, respectively.

**Table 1.** Characteristics of 27 critically ill COVID-19 patients according to the clinical classification of aspergillosis. Comparisons were performed using Chi-2 or Mann-Whitney tests, as appropriate.

| Parameters | Putative aspergillosis<br>(n=9) | No aspergillosis<br>(n=18) | p |
| --- | --- | --- | --- |
| Age (years) | 63 | 63 | 1.0 |
| Sex ratio (M/F) | 1.3 | 2.6 | 0.4 |
| Diabetes, % | 33 | 33 | 1.0 |
| Hypertension, % | 78 | 33 | **0.04** |
| Obesity, % | 33 | 17 | 0.4 |
| Ischemic heart disease, % | 22 | 33 | 0.7 |
| Aspergillosis host factors, %† | 22 | 17 | 1.0 |
| Time from symptoms start to tracheal intubation (days) | 7 [3-10] | 8 [5-13] | 0.2 |
| Time from tracheal intubation to *Aspergillus* pulmonary specimen (days) | 6 [1-11] | 2 [0-10] | 0.1 |
| Time from symptoms start to *Aspergillus* pulmonary specimen (days) | 12 [7-15] | 11 [5-21] | 0.7 |
| Viral load, (cycles) | 24 [17-37] | 27 [15-34] | 1.0 |
| PaO<sub>2</sub>/FiO<sub>2</sub> (mmHg) | 167 [88-290] | 163 [42-480] | 0.8 |
| Non-COVID CT scan signs, % | 67 | 43 | 1.0 |
| BAL macrophages, % (N=20) | 26 [8-55] | 34 [4-49] | 0.8 |
| BAL neutrophils, % (N=20) | 47 [10-82] | 30 [13-87] | 1.0 |
| BAL lymphocytes, % (N=20) | 12 [6-48] | 24 [1-53] | 0.6 |
| Vasopressors, % | 89 | 67 | 0.4 |
| Renal replacement, % | 30 | 22 | 0.7 |
| Steroid administration in the ICU, %†† | 67 | 72 | 1.0 |
| Deaths, % | 33 | 17 | 0.4 |

†based on EORTC MSG criteria for invasive aspergillosis, including steroids (n=3) and haematological malignancies (n=2); ††Steroid regimen, dexamethasone intravenous dose of 20 mg once daily from day 1 to day 5, followed by 10 mg once daily from day 6 to day 10; BAL, bronchoalveolar lavage; ICU, intensive care unit; CT scan, computerized tomography scanner.

Here we found putative IPA in almost one third of our successive critically ill COVID-19 patients, at a similar rate to what has been observed in influenza patients.^3,4^ Interestingly, when respiratory specimens were positive for *Aspergillus*, serum galactomannan was negative. One case had positive serum beta-D-glucan and galactomannan without *Aspergillus* detection in the BAL. Our findings strongly suggest that mechanically ventilated COVID-19 patients should be systematically screened for *Aspergillus* infection markers. Three of our cases had *Aspergillus fumigatu*s culture with no positive qPCR detection or galactomannan antigen in the BAL. Interestingly, not considering positive culture alone as a diagnostic criterion, by contrast to what is currently accepted,4,6 would have resulted in underestimating the frequency of putative IPA (22% rather than 33% in our study).

Despite similar IPA rates in critically ill COVID-19 and influenza patients, the contribution of Aspergillus to the patient presentation in each illnesses may be different. Here, none of our nine putative IPA patients except one received anti-*Aspergillus* treatment. In the treated patient, antifungal treatment was adapted to cover both candida and *Aspergillus* infections. In the IPA group, the three fatalities were not related to aspergillosis but to bacterial septic shock complicated by multiorgan failure.

Finally, although oseltamivir-induced inhibition of the host neuraminidase activity has been suggested as a possible molecular mechanism leading to anti-aspergillus protective immunity decrease in influenza patients, the exact reasons for increased vulnerability of the COVID-19 patient to *Aspergillus* remain to be determined as well as the *Aspergillus* contribution to COVID-19-reated lung inflammation.

## Data Availability

Data will be available upon publication

## Ethics

This study was part of the French COVID-19 cohort registry conducted by the REACTing consortium and directed by INSERM and ISARIC. Our institutional ethics committee approved the study (N°, IDRCB, 2020-A00256-33; CPP, 11-20 20.02.04.68737). When possible, signed informed consent was obtained from the patients or the next of kin.

## References

1 Wu Z, McGoogan JM. Characteristics of and Important Lessons From the Coronavirus Disease 2019 (COVID-19) Outbreak in China: Summary of a Report of 72-314 Cases From the Chinese Center for Disease Control and Prevention. JAMA 2020. DOI:10.1001/jama.2020.2648.

2 Lescure F-X, Bouadma L, Nguyen D, et al. Clinical and virological data of the first cases of COVID-19 in Europe: a case series. Lancet Infect Dis 2020. DOI:10.1016/s1473-3099(20)30200-0.

3 Wauters J, Baar I, Meersseman P, et al. Invasive pulmonary aspergillosis is a frequent complication of critically ill H1N1 patients: a retrospective study. Intens Care Med 2012; 38: 1761–8.

4 Schauwvlieghe AFAD, Rijnders BJA, Philips N, et al. Invasive aspergillosis in patients admitted to the intensive care unit with severe influenza: a retrospective cohort study. Lancet Respir Medicine 2018; 6: 782–92.

5 Donnelly JP, Chen SC, Kauffman CA, et al. Revision and Update of the Consensus Definitions of Invasive Fungal Disease From the European Organization for Research and Treatment of Cancer and the Mycoses Study Group Education and Research Consortium. Clin Infect Dis Official Publ Infect Dis Soc Am 2019. DOI:10.1093/cid/ciz1008.

6 Blot SI, Taccone FS, Abeele A-MV den, et al. A Clinical Algorithm to Diagnose Invasive Pulmonary Aspergillosis in Critically Ill Patients. Am J Resp Crit Care 2012; 186: 56–64.

7 Alanio A, Menotti J, Gits-Muselli M, et al. Circulating Aspergillus fumigatus DNA Is Quantitatively Correlated to Galactomannan in Serum. Front Microbiol 2017; 8: 2040.

